# *In vivo* evaluation of the virucidal efficacy of Chlorhexidine and Povidone-iodine mouthwashes against salivary SARS-CoV-2

**DOI:** 10.1101/2021.03.07.21252302

**Authors:** Rola Elzein, Fadi Abdel-Sater, Soha Fakhreddine, Pierre Abi Hanna, Rita Feghali, Hassan Hamad, Fouad Ayoub

## Abstract

**Background:** The oral cavity is potentially high-risk transmitter of COVID-19. Antimicrobial mouthrinses are used in many clinical pre procedural situations for prophylactic purposes. An evident-based investigation for an effective mouthwash solution against salivary SARS-CoV-2 is urgently required for the exposure reduction during dental procedures.

**Aims:** This study aimed to evaluate in vivo virucidal efficacy of 2 mouthwashes: 1% Povidoneiodine and 0.2% Chlorhexidine as a dental preprocedural oral disinfection against salivary SARS-CoV-2.

**Materials and Methods:** In this randomized-controlled clinical trial, studied group comprised laboratory-confirmed COVID-19 positive patients through nasopharyngeal swabs. Participants were divided into 3 groups. For 30 seconds, group A gargled with 1% Povidone-iodine, group B mouthrinsed with 0.2% Chlorhexidine and control group C mouthrinsed with distilled water. Saliva samples were collected before and 5 minutes after mouthwash. SARS-CoV-2 rRT-PCR was then performed for each sample. Evaluation of the efficacy was based on difference in Ct value. The analysis of data was carried out using *GraphPad Prism* version 5 for Windows. Paired t test and unpaired t test were used. A probability value of less than 0.05 was regarded as statistically significant.

**Results:** Sixty-one compliant participants (36 female and 25 male) with a mean age 45.3 ± 16.7 years-old were enrolled. A significant mean Ct value difference (*p* < 0.0001) between the paired samples in group A (n = 25) and also in group B (n = 27) (*p* < 0.0001) was found. In contrast, no significant difference (*p* = 0.566) existed before and after the experiment in the control group C (n = 9). Moreover, a significant difference was noted between the delta Ct of distilled water wash and each of the 2 solutions 1 % Povidone-iodine (*p* = 0.012) and Chlorhexidine 0.2% (*p* = 0.0024). No significant difference was found between the delta Ct of patients using 1% Povidone-iodine and Chlorhexidine 0.2% solutions (*p* = 0.24).

**Conclusion:** Chlorhexidine 0.2% and 1% Povidone-iodine oral solutions are effective preprocedural mouthwashes against salivary SARS-COV-2 in dental treatments. Their use as a preventive strategy to reduce the spread of COVID-19 during dental practice should be systematically implemented.

## Introduction

Severe acute respiratory syndrome coronavirus 2 (SARS-CoV-2) is the novel member of the human coronaviruses from the *Coronaviridae* family and belongs to the *Betacoronavirus* genus.^1^ The subsequent corona virus disease-2019 (COVID-19), rapidly spreading worldwide,^2^ mostly causes respiratory disorders.^3,4^ The human-to-human transmission of SARS-CoV-2 essentially happens by inhalation of respiratory droplets spread by coughing or sneezing from an infected person, and by direct contact of contaminated surfaces followed by touching the nose, mouth and eyes.^5,6^ The virus can even survive on various surfaces for days.^7^ Transmission via ocular has been shown.^8^ The oral cavity is potentially high-risk transmitter of COVID-19. In fact, when operating dental treatments with the high-speed handpiece, it is fundamental to use a water coolant,^9^ generating consequently aerosols mixed with saliva or blood. These bioaerosols, generally contaminated with microorganisms including bacteria, fungi, and viruses, float in the air then settle on the surfaces and can be transmitted to the dentists or other patients by inhalation or contact.^10,11^ SARS-CoV-2, was identified in saliva of infected patients.^12^ Furthermore, it has been reported that the main cell receptor of SARS-CoV-2, angiotensin-converting enzyme II (ACE2), is extremely expressed on the mucosa of the oral cavity and particularly in the epithelial cells of the tongue.^13^ Therefore, it is crucial for dental practitioners to install preventive strategies to avoid the COVID-19 infection by focusing not only on patient placement, hand hygiene, all personal protective equipment, caution in performing aerosol-generating procedures but also on patient’s pre-procedural antiseptic mouthrinse.^14,15^ In fact, antimicrobial mouthrinses are an important part of oral care. Such solutions are used in many clinical pre procedural situations for prophylactic purposes.^16,17^ Preprocedural oral solution is one of the most effective methods of reducing the amount of microorganisms in oral aerosols.^18,19^ In addition, gargling is also assumed to produce favorable effects through removal of oral and pharyngeal protease that helps viral replication.^20^

Thus, an investigation for an effective mouthrinse against COVID-19 is urgently required for the control of oral and respiratory tract infection and for the exposure reduction during dental procedures.

In the literature, it was reported that a pre-procedural 0.12% Chlorhexidine mouth rinse can reduce the microbial load of saliva.^21^ A meta-analysis showed that the use of preprocedural mouth rinse, including Chlorhexidine, essential oils, and cetylpyridinium chloride, resulted in a mean reduction of 68.4% colony-forming units in dental aerosol.^22^ Although the effect of Chlorhexidine gluconate on human coronavirus is unknown but it is effective against many respiratory viruses, like herpes and HIV.^23^ On the other hand, Povidone-iodine is a broad-spectrum antimicrobial that has been used in infection control and prevention for over 60 years and is available in various preparations for use as a disinfectant for the skin, hands and mucosal surfaces, as well as for wound treatment and eye applications.^24^ Povidone-iodine has well-established general antimicrobial activity, demonstrating *in vitro* efficacy against wide range of enveloped and nonenveloped viruses.^25-27^ Recent in vitro study has demonstrated rapid virucidal its products activity against MERS-CoV.^28,29^ The benefit of gargling with Povidone-iodine has also already been noted in Japanese clinical respiratory guidelines.^30^

This study, besides of being an additional research concerning the Consistency of detection of SARS-CoV-2 in saliva from the Lebanese experience, aimed mainly to evaluate *in vivo* virucidal efficacy of 2 mouthrinses: 1% Povidone-iodine and 0.2% Chlorhexidine as a dental preprocedural oral disinfection against salivary SARS-CoV-2.

## Materials and methods

### Ethical approval

Ethical clear of this research was delivered from the Lebanese University Institutional Review Board (#CUER 13-2020). The study was conducted in accordance with the 1964 Helsinki declaration and its later amendments or comparable ethical standards. All participants were informed about the study and gave their consent. It was conducted between June and Septembre 2020 at the isolation ward of Rafik Hariri University Hospital (RHUH) of Beirut for sampling and in the Laboratory of Cancer Biology and Cellular Immunology, COVID-19 Unit, Faculty of Sciences, Lebanese University for viral PCR tests.

### Trial design

This study was a parallel group, quadruple blind-randomized-placebo-controlled clinical trial with an add on laboratory based study. Patients, intervention supervisor, laboratory technicians and the data collection subject were blinded. The groups were labelled as A, B and C. The codes of the intervention were only revealed at the end of study.

### Participants

A non-probability, purposive sampling technique was adopted. Studied group comprised, laboratory-confirmed COVID-19 positive patients, through nasopharyngeal swabs. The lapse of time between COVID-19 diagnosis and inclusion in the study ranged from zero to two days. Were excluded cases indicated for intubation or mechanical ventilation and patients who declined consent.

### Solution preparation

15 ml of each undiluted mouthwash solution were previously poured into a sterile cup within a Biosafety cabinet (Topair system) in the Laboratory of Cancer Biology and Cellular Immunology, Faculty of Sciences, Lebanese University with respect to the manufacturer recommendations for each solution. The containers were marked as A, B or C, and then delivered to RHUH to accomplish the sampling. Solution A referred to 1% Povidone-iodine, B for 0.2% Chlorhexidine and C for distilled water as a placebo treatment.

### Sampling

Participants were randomly divided into the three groups. The same trained operator explained, provided and supervised the sampling in patient’s room with respect to COVID-19 infection control. Sampling was performed by the patients themselves in the early morning on empty stomach and before brushing teeth. First, participants were asked to cough out saliva from throat (2ml), into a first sterile container. Next, group A participants (n = 33) were invited to gargle for 30 seconds with solution A, group B (n = 33) to mouthrinse for 30 seconds with solution B and group C (n = 11) to mouthrinse for 30 seconds with solution C and then to spit the solution. Five minutes later, saliva collections were done again in a second sterile container. Each cup held patient’s name and the date of saliva collection while contaminated waste was appropriately discarded. Each collected sample was then inserted into separated tubes containing 2 mL of the virus transport medium (VTM) and transported to the COVID unit Laboratory in the Lebanese university for PCR processing.

### SARS-CoV-2 rRT-PCR

The presence of SARS-CoV-2 was confirmed by real-time reverse transcriptase polymerase chain reaction (RT-PCR). 200 µL of VTM was used for RNA purification. RNA was extracted from the clinical samples on Kingfisher flex purification system Thermo Fisher using MagMAX(tm) Viral/Pathogen Nucleic Acid Isolation Kit (thermos fisher). Reactions were performed in 20 µL final volume reaction containing 5 µL of extracted RNA, rRT-PCR was performed using CFX96 real-time PCR detection system (Bio-Rad, Hercules, CA, USA) and Bosphore Novel Coronavirus (2019-nCoV) PCR Detection Kit v4 (Anatolia, Turkey), which targeted the RdRP, N and E genes of SARS-CoV-2. In this assay, a RNase P gene region is used as an endogenous internal control for the analysis of biological samples. It is normally used to ensure the quality of the test, at extraction and PCR levels and to exclude the false negative results. Thus, in order to evaluate possible variability in the amount of material retrieved from saliva specimen before and after mouth wash we utilized RNase P as reference gene to normalize the input data.

To compare the paired samples before and after mouth wash, we calculated a Ct value modified according to the ratio of sample RNase P and mean RNase P Ct values.^31^

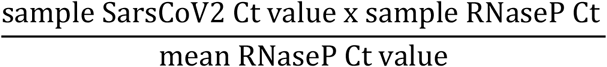

### Statistical Analysis

The analysis of data was carried out using *GraphPad Prism* version 5 for Windows (*GraphPad* Software, La Jolla California USA, www.graphpad.com). Differences between the means of the two matched groups were explored using Paired t test. The means of the two independent Groups were compared by using the Unpaired t test. A probability value of less than 0.05 was regarded as statistically significant.

## Results

In total, while 77 patients were eligible for the study, 16 were excluded resulting in 61 compliant participants (Figure 1). Among the final study group, 36 (59.1%) were female and 25 (40.9%) were male. The mean age of all patients was 45.3 ± 16.7 with an age range between 17 and 85 years old. The description of each group is mentioned in table 1.

**Table 1.** Description of the study population.

| Participants | Age years |  | Gender |  |
| --- | --- | --- | --- | --- |
| | Median (range)<br>(years) | Mean ± SD<br>(years) | Female<br>N(%) | Male |
| All patients<br>N = 61 | 43 (17-85) | 45.3 ± 16.7 | 36 (59.1%) | 25 (40.9%) |
| Distilled water group<br>N = 9 | 56 (21-85) | 57.2 ± 22.5 | 7 (77.8%) | 2 (22.2%) |
| Povidone iodine 1% group<br>N = 27 | 40 (17-63) | 39.9 ± 14.2 | 14 (51.8%) | 13 (48.2%) |
| Chlorhexidine 0.2% group<br>N = 25 | 51 (25-84) | 47 ± 15.4 | 15 (60%) | 10 (40%) |

**Figure 1.**
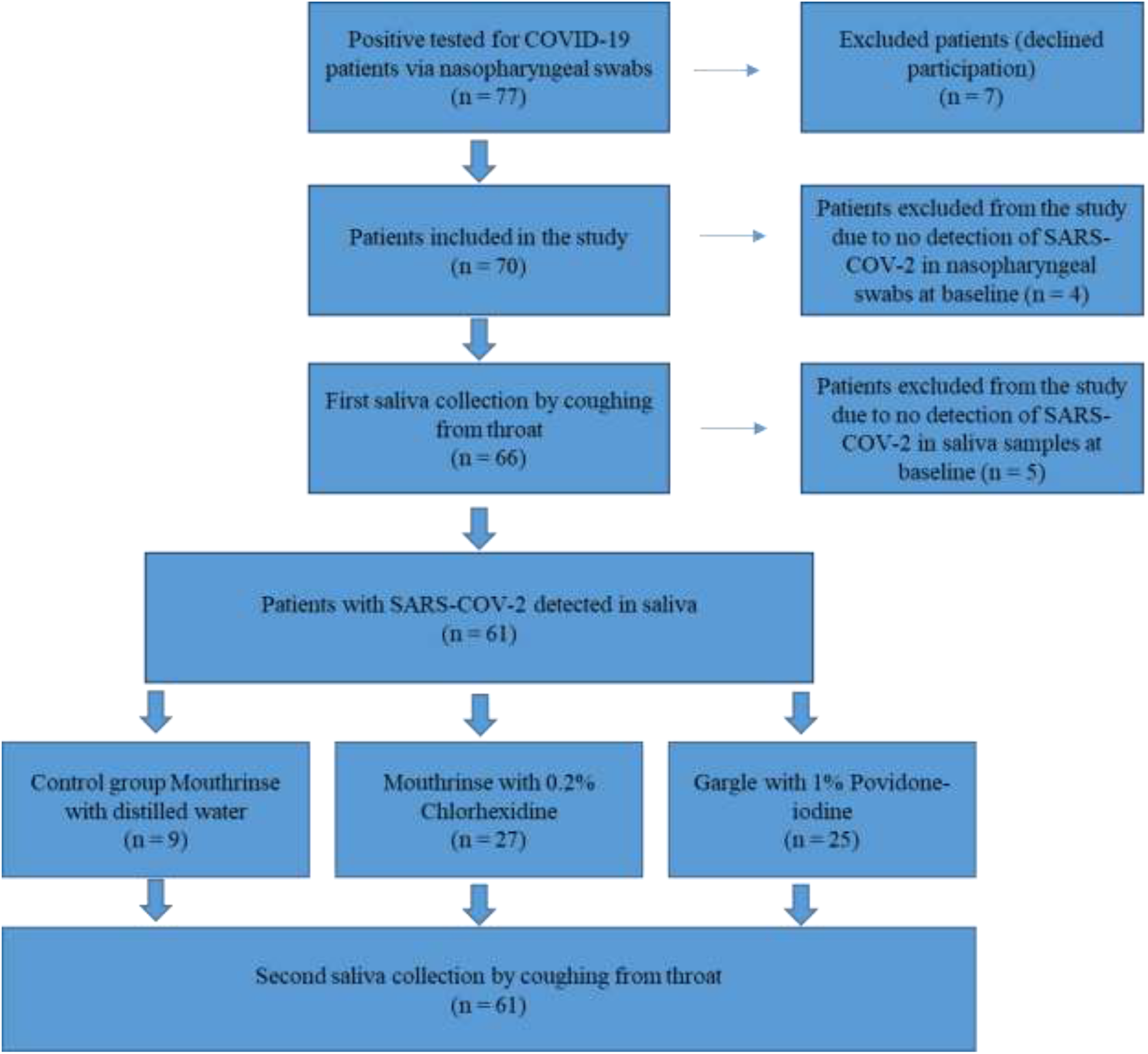
Flow diagram showing the enrolment process of participants in the study.

The mean Ct value of human RNaseP in saliva samples before mouthwash was 25.41 ± 2.5 [18.4– 32.21]. Among the specimens tested, 72.4% had RNaseP Ct values below 27, 20.7% between 27 and 30, and 7.9% between 30 and 32.2. The mean Ct value of human RNaseP in saliva samples after mouthwash was 26 ± 2.72 [19.49-32.5]. No significant difference was found between the mean Ct values of human RNaseP in the 2 groups (*p* = 0.332).

The expression of the SARS-CoV-2 target genes (RdRp, E and N) used was approximately the same in each tested sample. To simplify our analysis, we presented the results with RdRp. For this gene the mean Ct value was 28.9 ± 5.5 (Median 29.9 [16.45–38.16]) in salivary pre-wash samples. After normalization, the SARS-CoV-2 mean Ct value was 28.3 ± 6.3.

Our results showed a significant mean difference between the paired samples before (29.88 ± 6.2; median 30.75) and after mouthwash (34.36 ± 6.3; median 34.19) with 1% Povidone-iodine (*p* < 0.0001). After mouthwash, the difference between means was 4.45 (Figure 2A). In addition, a higher significant difference of means was found in paired samples using Chlorhexidine 0.2% (*p* < 0.0001). The mean Ct increased 5.69 after mouthwash. The mean Ct of pre and post mouthwash was respectively 27.69 ± 7.16 (median 27.11) and 33.9 ± 7.08 (median 33.13) (Figure 2B). In contrast, no significant difference was found in the control group using the distilled water as mouthwash solution as shown in figure 2C (*p* = 0.566). Moreover, the comparison of the delta Ct showed a significant difference between distilled water wash and each of the 2 solutions Povidone-iodine and Chlorhexidine 0.2% (*p* value 0.012 and 0.0024 respectively). No significant difference was found between the delta Ct of patients using 1% Povidone-iodine and Chlorhexidine 0.2% solutions (*p* = 0.24) (Figure 3). We noted that Ct values are considered inversely related to viral load and may serve as an indirect method of arbitrarily quantifying the viral load in the sample.

**Figure 2.**
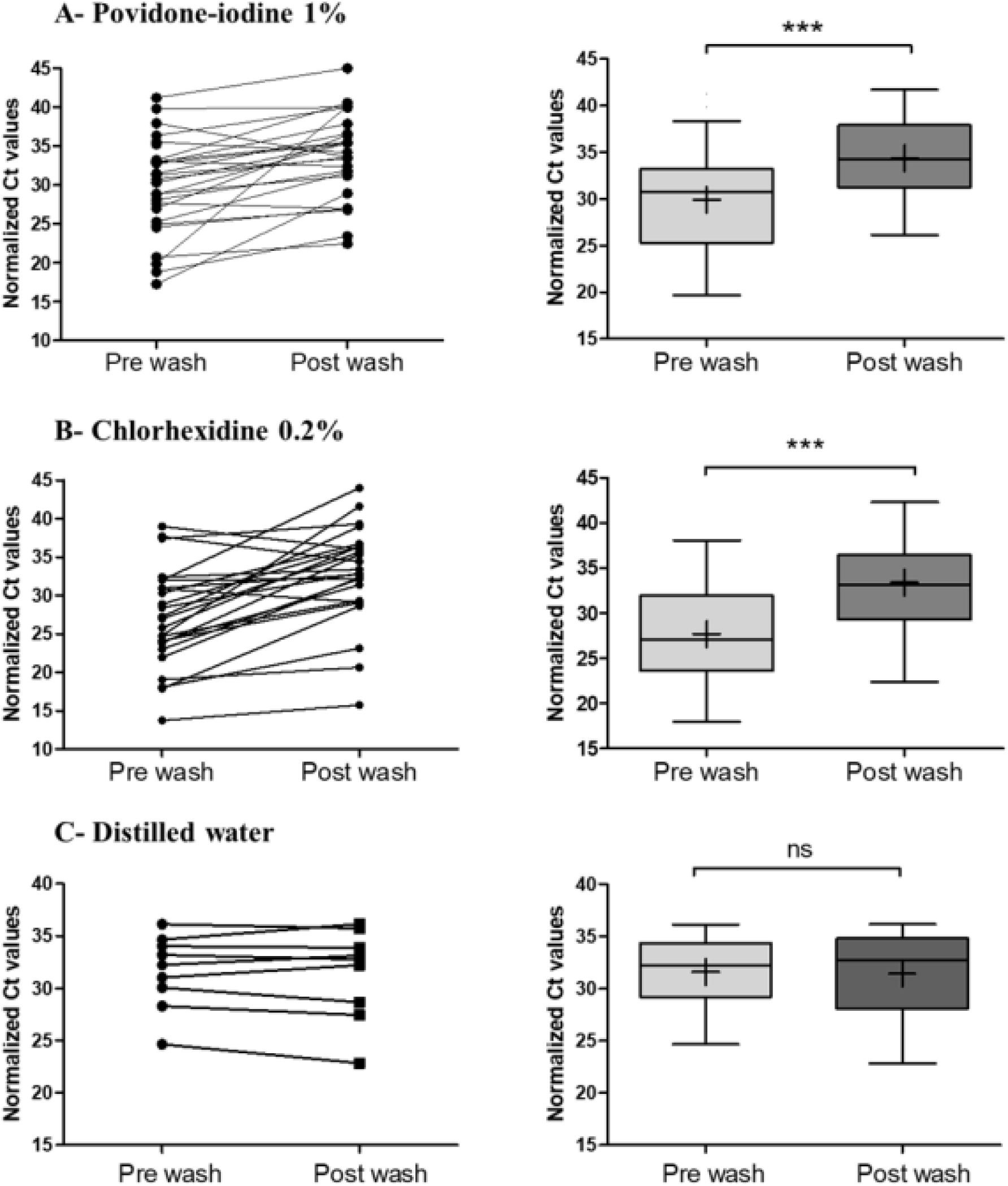
Normalized threshold cycle (Ct) values for matched pre and post mouthwash salivary specimens. The Ct values of *RdRp* obtained with RT–PCR that were detected in salivary specimens before and after mouthwash and normalized to the internal control RNAseP endogenous human gene. (A) 1% Povidone-iodine (n = 25), (B) Chlorhexidine 0.2% (n = 27) and (C) distilled water (n = 9) were used as mouthwash solutions. The before-after graph shows the changes of Ct after mouth wash for each patient. The box plots show the medians (middle line) and the first and third quartiles (boxes). The mean is marked by a plus sign inside the box. Paired Groups were compared by using the Paired t test. ***Indicates a p-value ≤ 0.0001, ns indicates no significant difference (*p* > 0.05).

**Figure 3.**
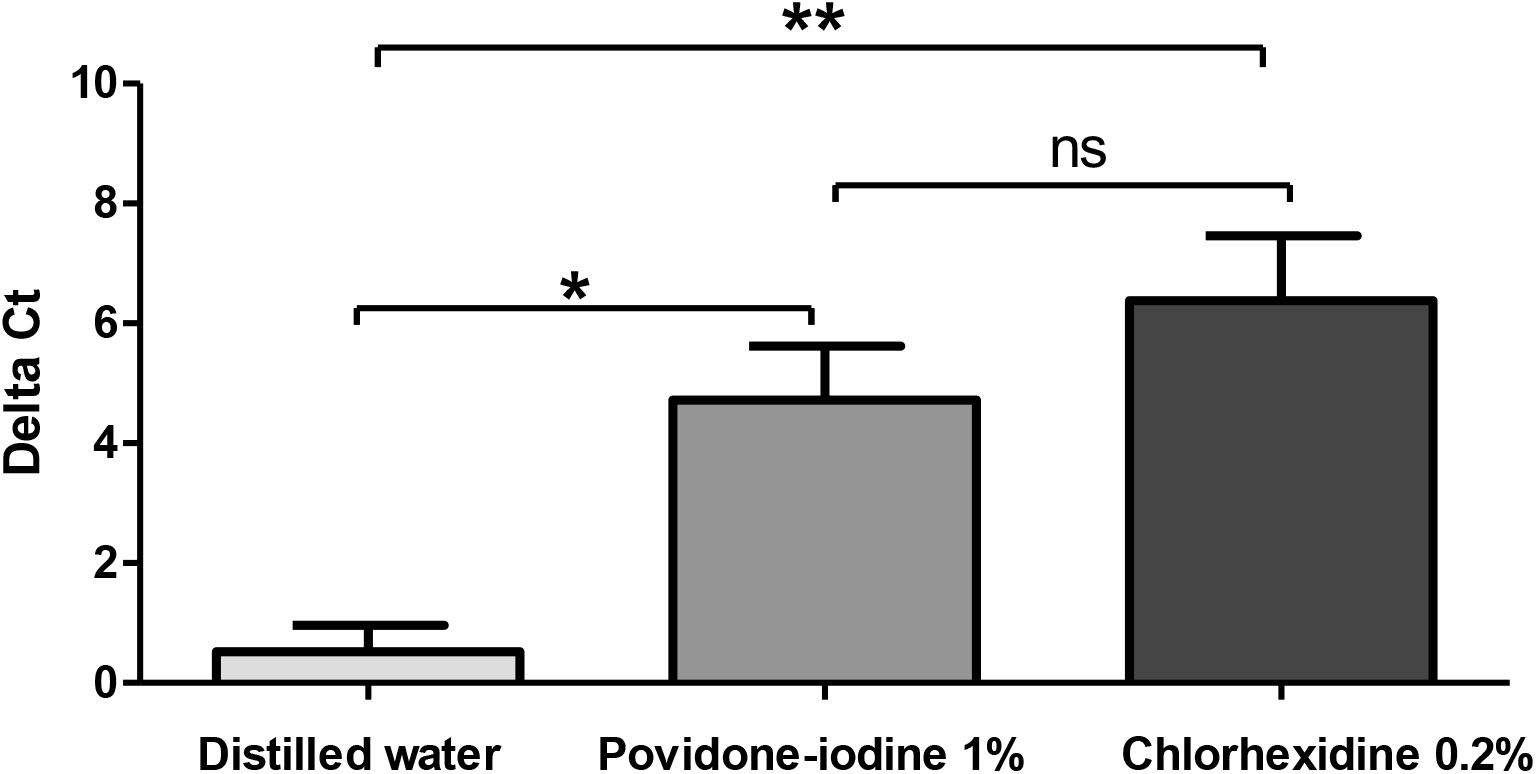
Comparison of Delta Ct mean between the mouth wash solutions. Delta Ct were calculated as follow: normalized Ct value post mouthwash minus normalized Ct value pre mouthwash. Groups were compared by using the Unpaired t test. Peak values are reported as mean +/-SD. Significant differences between means are indicated by *(*p* < 0.05), ** (*p* < 0.01), ns indicates no significant difference (*p* > 0.05).

## Discussion

With the novel COVID-19 pandemic, dental care practitioners were in urge to develop quick infection control policies.^32^ So far, clinical implementation of new concepts was mainly based on recommendations without being evident based. Particularly, different pre-procedural mouthwash solutions to minimize the SARS-CoV-2 transmission during dental treatment were recommended by some dental specialist societies.^33-35^ Despite lack of any clinical data supporting the virucidal effects of mouthwash solutions against SARS-CoV-2, many propositions were adopted in reviews discussing the COVID-19 preventive measurements in Dentistry.^36-41^ A recent in-vitro study tested the effect of the following mouth rinses on cell viability: hydrogen peroxide, povidone-iodine, chlorhexidine gluconate and essential oils with alcohol. The experiments found that mouth rinses can significantly reduce virus infectivity, suggesting a potential benefit for reducing SARS-CoV-2 spread. The study concluded that the clinical investigation of antiviral effects of mouth rinses is needed for proving their potential to reduce the virus spread.^42^

For our knowledge this current *in vivo* study is the first large scale controlled-clinical trial testing the efficacy of 1% Povidone-iodine oral gargle and 0.2% Chlorhexidine oral mouthwash on salivary SARS-CoV-2 virus of positive tested patients.

This study comprised 61 compliant COVID-19 positive subjects. During sampling recruitment, the non-detection of SARS-CoV-2 in nasopharyngeal samples of four hospitalized positive patients could be explained by the fact that the nucleic acid test results of a significant proportion of patients are “false negative”.^43^ For ethical issues, mainly avoiding the patient subsequent discomfort, we preferred to exclude these participants instead of repeating the nasopharyngeal swab test for a clinical trial purpose.

In placebo-controlled clinical trials with “very ill” subjects it is unethical to assign equal subjects to each arm and it is preferable to have more subjects in test group compared to the control one. In such cases, sample size is adjusted if clear and clinically meaningful inputs on some points are available prior to working on sample size estimation.^44^ As COVID-19 is a recently emerging pandemic without previous related data in addition to its critical incompletely explored status, we considered that our sample size for the test and control groups were legible.

Saliva sampling was self-performed by the patients to reduce the risk of nosocomial SARS-CoV-2 transmission to health care providers.^12,45^ A lapse of time of 5 minutes between mouthrinsing or gargling and second saliva collection was chosen conforming to the real procedure at dental clinic: during dental appointment, it takes usually few minutes between the patient’s preprocedural mouthwash and the commencement of the treatment.

This study firstly revealed the consistency of the detection of SARS-CoV-2 in saliva since the virus was detected in the salivary samples of 61 out of 66 patients. This result is in accordance with other studies.^12,46,47^ In fact, using saliva specimens for the diagnosis of COVID-19 has many advantages like avoiding invasive procedures, contributing to the decrease of the risk of nosocomial COVID-19 transmission, usefulness for screening of a large number of individuals with less time consuming and in situations in which nasopharyngeal specimen collection may be contraindicated.^12^ However, further studies comparing SARS-CoV-2 viral load between saliva and nasopharyngeal samples collected at the same time for each COVID-19 tested patient are required for a better assessment for the use of saliva as a diagnosis tool for COVID-19.

Moreover, our results showed that a 1% Povidone-iodine gargle reduces significantly the intraoral viral load in SARS-CoV-2-positive subjects. Our results are similar to another in-vivo study where the authors analyzed the impact of a mouthwash with Povidone-iodine on the salivary viral load of SARS-CoV-2 in 4 patients with COVID-19 and found that in 2 of the 4 participants, the gargle solution resulted in a significant drop in viral load, which remained for at least 3 h.^48^ Frank *et al*. (2020) found that Povidone-iodine can safely be used in the mouth at concentrations up to 2.5% for up to 5 months because it rapidly inactivates coronaviruses, including SARS and MERS, even when applied for as little as 15 seconds.^49^ The same authors were optimistic about the inactivation of SARS-CoV-2 by Povidone-iodine, and called for in vitro efficacy demonstration. Mady *et al*. (2020) proposed the use of oral/oropharyngeal wash with 10 mL of 0.5% aqueous Povidone-iodine solution in addition to nasal irrigation of 240 mL of 0.4% of the same antiseptic solution for patients and healthcare providers as a public health intervention for COVID-19.^50^ Suresh *et al*. (2020) proposed also a pioneer description in anesthesia practice on the use of preoperative Povidone-iodine gargles in COVID-19 cases to mitigate the chain of spread of COVID-19 through cross-infection among health care workers.^51^ In addition, Brida *et al*. (2020) investigated the in-vitro optimal contact time and concentration for virucidal activity of 0.5%, 1%, and 1.5% oral solution of povidone-iodine against SARS-CoV-2 and found that the efficacy was present at the lowest concentration of 0.5 % Povidone-iodine and at the lowest contact time of 15 seconds and mentioned that, therefore, preprocedural rinsing with diluted Povidone-iodine in the range of 0.5% to 1.5% may be preferred over hydrogen peroxide during the COVID-19 pandemic.^52^ Pelletier *et al*. (2020) found that concentrations 1% to 5% of Povidone-iodine nasal antiseptics and oral rinse antiseptics completely inactivated the SARS-CoV-2 after 60-second exposure times on SARS-CoV-2 infected Vero 76 cell.^53^ On the other hand, Chorney *et al*. (2020) called for further research prior to strongly recommending Povidone-iodine use in preparation for nasal, oral or pharyngeal surgery in children.^54^

In addition, preprocedural mouthrinse with 0.2% Chlorhexidine showed in our study a significant efficacy against SARS-CoV-2. Our results are in accordance with those of Yoon *et al*. (2020) who found, in a clinical trial on 2 patients, that Chlorhexidine mouthwash was effective in reducing the SARS-CoV-2 viral load in the saliva for a short-term period.^55^ Meister *et al*. (2020) found while using Vero E6 cells that different SARS-CoV-2 strains can be efficiently inactivated with Chlorhexidine and other commercially available oral rinses and recommended further analysis during clinical studies to assess the in vivo effects of the oral solutions. Chlorhexidine has been suggested to reduce the viral transmission via aerosols.^56^ Although its action against this virus remains controversial but if the results are confirmed by other clinical trials, Chlorhexidine mouthrinse could help to prevent the spread of SARS-CoV-2.^57^

Although both solutions proved significant efficacy against salivary SARS-CoV-2, 0.2% Chlorhexidine showed non-significantly more efficiency on reducing the salivary viral load than 1% povidone iodine. Distilled water had no effect on viral load. The absence of placebo effect confirmed the effectiveness of the proposed disinfectant mouthwash solutions on salivary SARS-COV-2.

## Conclusion

0.2% Chlorhexidine and 1% Povidone-iodine oral solutions are effective preprocedural mouthwashes against SARS-COV-2 in dental treatments. Their use as a preventive strategy to reduce the spread of COVID-19 should be systematically implemented in Dentistry as in various health care services. Further studies including the length of their effectiveness over the time are required for an accurate prescription against SARS-COV-2.

## Data Availability

Data available upon request

